# Programmable kinetic barcoding for multiplexed RNA detection with Cas13a

**DOI:** 10.1101/2025.07.03.25328829

**Authors:** Sungmin Son, Amy Lyden, Carlos F. Ng, Andres Dextre, Jeffrey Shu, Stephanie I. Stephens, Parinaz Fozouni, Gavin J. Knott, Dylan C. J. Smock, Tina Y. Liu, Daniela Boehm, Camille Simoneau, G. Renuka Kumar, Jennifer A. Doudna, Melanie Ott, Daniel A. Fletcher

## Abstract

Rapid identification of viral infections and specific variants in patient samples requires a simple and multiplexed RNA detection method that does not rely on DNA sequencing. Although recent direct detection assays based on CRISPR-Cas13a^1–4^ offer rapid RNA detection by avoiding reverse transcription and DNA amplification required of gold-standard PCR assays^5^, these assays are not easily multiplexed to detect multiple viruses or variants without dividing the sample into separate reactions. Here we show that Cas13a acting on single target RNAs exhibits variable nuclease activity that depends on the interaction between the target RNA and crRNA. To exploit this feature for multiplexed detection, we devised a crRNA modification strategy that enables programmable tuning of Cas13a’s nuclease enzymatic rates. Using a droplet-based Cas13a assay, we demonstrate that kinetic signatures can be harnessed to differentiate among respiratory viruses and SARS-CoV-2 variants in contrived and clinical samples. This kinetic barcoding strategy can be extended to additional RNA targets through simple modification of crRNAs.

## MAIN TEXT

Diagnostic testing for viral infections must be rapid and specific in order to accurately monitor the spread of variants and enable timely responses. In the case of SARS-CoV-2, where symptoms are non-specific and often indistinguishable from other respiratory infections, accurate identification of virus is crucial for optimal and timely treatment^6^. While genomic sequencing provides the most complete information about a virus, a more rapid diagnostic method that can simultaneously detect multiple viral targets and variants is also needed. Although commercial platforms are available that offer multiplexed detection based on PCR reactions^7,8^, they typically require complex hardware and long times. CRSIPR-Cas systems such as Cas13a offer an alternative method for detecting nucleic acids based on sequence-specific activation^9^. Though Cas13 can be combined with reverse transcription, DNA amplification, and transcription to increase sensitivity^10,11^, we previously demonstrated that Cas13 can avoid limitations associated with those steps and sensitively detect viral RNA without amplification of its gene^1^.

Here we show that Cas13-based RNA detection can be multiplexed by quantifying differences in single enzyme kinetics with a droplet-based version of the Cas13 direct detection assay, where droplet size makes it likely for a maximum of one target RNA to end up in each reaction. This strategy, which we call ‘kinetic barcoding’, arises from the observation that Cas13 complexed with a single target RNA exhibits variable nuclease activity depending on the specific crRNA and target RNA present. This variability is well known from guide screening efforts necessary to identify a subset of ‘good’ crRNA with high nuclease activity for a given target RNA. We find that such natural differences in kinetics are sufficient to distinguish different viral targets in the droplet Cas13 assay. Moreover, we show that the average enzymatic rate of Cas13 nuclease can be altered in a controlled way by adding single-stranded DNA to the 5’ end of the crRNA. Addition of 10 or more deoxynucleotides to the crRNA results in a reproducible and quantifiable stepwise decrease in cleavage rates of fluorophore-quencher molecules used as reporters. By combining multiple crRNAs targeting different viruses or variants that have been modified through the addition of a 5’ DNA sequence to produce distinct activity, the identity of a single target RNA can be determined solely based on the slope or endpoint value of the fluorescence signal, without the need for separate fluorescence channels. In contrast to other multiplexing strategies that divide samples and then look for different RNA targets in each reaction, kinetic barcoding simultaneously looks for all different RNA targets in all reactions since Cas13 and the barcoded crRNAs are present in all droplets, allowing assay multiplexing without any modifications in the assay workflow or in the hardware.

### A droplet Cas13 assay enables single enzyme kinetics measurements

We first developed a simple and rapid droplet Cas13 assay using emulsification. The small droplet volumes accelerate the accumulation of cleaved fluorophores from reporter molecules (Fig. 1A) in addition to encapsulating single target RNAs, making it possible to precisely measure single Cas13 enzyme activity. We generated approximately 10 pL droplets by emulsifying a reaction mix containing target RNA, crRNA, fluorophore-quencher reporter, and LbuCas13a for 2 min in an excess volume of an oil/surfactant/detergent mixture using an automatic multichannel pipettor and a pipetting robot (Extended Data Fig. 1A, Methods, Supplementary Video 1). We loaded the resulting emulsion, with diameters ranging from 10 to 40 µm, into a custom imaging chamber and directly imaged the reaction in time on a fluorescence microscope (Fig. 1B, Extended Data Fig. 1B and 1C). Imaging the droplets allowed us to normalize the fluorescence signal by droplet size^12^, avoiding the need for slower and low throughput systems for generating uniform droplets.

**Figure 1:**
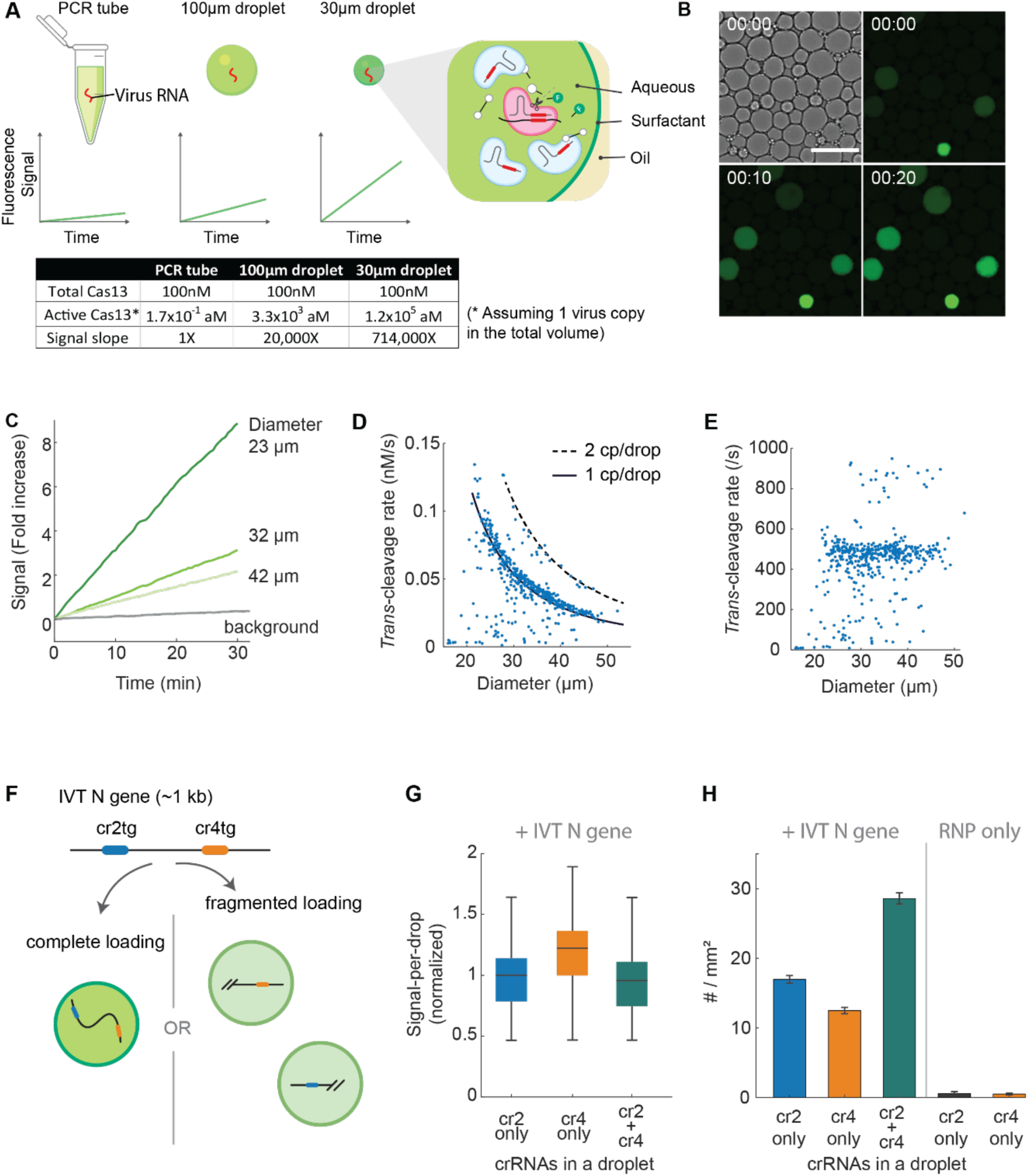
A droplet assay measures Cas13a single molecule kinetics. a) Schematic showing the increased signal accumulation rate for a single Cas13a confined in decreasing volumes (red: target RNA). The table quantitatively shows the effect of reaction volume on the signal slope, assuming only a single target is present. Each droplet contains hundreds of thousands copies of the Cas13a-crRNA RNP (ribonucleoprotein complex) and millions of quenched RNA reporters. b) Brightfield (top left) and fluorescence images of droplet Cas13a reactions taken with a 20X objective lens. T = 0, 10, and 20 minutes since the beginning of imaging. Scale bar = 65 µm. c) Fluorescence signal over time in three positive droplets (green lines) and one negative droplet (grey line). The signal is calculated from the mean fluorescent intensity change within a droplet normalized by its initial value after background subtraction. An image is taken every 30 seconds and corrected for photobleaching (Methods). d) Single Cas13a *trans*-cleavage rate measured from individual droplets containing crRNA 4, SARS-CoV-2 RNA, and 400 nM 5U-fluorescent reporter (N = 478 droplets). e) The data in d) is expressed in turnover-frequency by normalizing the signal for droplet size. f) Schematic showing two possible outcomes of a Cas13 droplet reaction combining two crRNAs. The RNA targets for crRNA2 and crRNA4 are cr2tg and cr4tg, respectively. If the whole N gene is loaded into a droplet containing both crRNAs, the fluorescence signal will accumulate twice as fast as the droplet containing only one target sequence (complete loading). However, if the N gene is fragmented prior to encapsulation and parts of the N gene are loaded into two different droplets, the number of positive droplets will be doubled, and the signal of each droplet will be the same (fragmented loading). g) The result of Cas13a reaction with 2.5 x 10^4^ copies/µL of IVT N gene and without any target RNA in droplets containing crRNA 2, crRNA 4, or both crRNAs are quantified after 1 hour of reaction incubation from three technical replicates. The distribution of signal-per-droplet is represented as a box and whisker plot showing the median, lower and upper quartiles, and minimum and the maximum values ignoring outliers. Data from the triplicate runs were combined for each condition. h) Same experiment as described in f), but the number of positive droplets is shown. The mean of total positive droplets is 14.8, 11.2, 25.6, 0.08, and 0.04, for crRNA 2, crRNA 4, or both crRNAs with target RNAs and crRNA 2 and crRNA 4 without any target, respectively. The mean and standard deviation were derived from triplicates.

Accurate determination of enzyme kinetics relies on knowing the active enzyme concentration. In bulk measurements of LbuCas13a kinetics^1^, it has been unknown what fraction of target RNA is actually activating LbuCas13a, leading to some debate^13^. In the droplet reaction, by contrast, the active enzyme concentration is known since the fluorescent signal is produced by a single target RNA activating LbuCas13a. We chose a crRNA targeting the SARS-CoV-2 N gene (crRNA 4) and formed droplets containing crRNA 4 along with SARS-CoV-2 RNA, LbuCas13a, and a fluorophore-quencher pair tethered by pentauridine (5U) RNA (reporter) (Fig. 1A). For a target RNA concentration of 1 x 10^4^ copies/µL, we found 7% of the droplets contain the target RNA, with more than 95% of those containing only a single copy, allowing direct measurement of a single-Cas13a enzyme kinetics. As expected, we found that the signal accumulation rate in positive droplets was inversely proportional to droplet size, with smaller droplets increasing faster than larger droplets (9-fold increase for a 23 µm droplet vs. 3-fold increase for a 42 µm droplet) (Fig. 1B, C, and D). We found that a single LbuCas13a can cleave 471 ± 47 copies of reporter every second in the presence of 400nM reporter (Fig. 1E), or a K_cat_/K_M_ of 1.2 x 10^9^ M^-1^s^-1^, which is two orders-of-magnitude higher than that measured for LbCas12a^14^ and consistent with that measured for LbuCas13a based in a bulk assay^15^ as well as in a digital assay^16^. This reveals that LbuCas13a is an exceptionally rapid nuclease, operating at the diffusion-limited regime. It is worth noting, however, that the actual enzyme kinetics using di-U reporter (2U) might be slower, as the addition of the poly-U reporter (5U) substantially elevates the local substrate concentration beyond 400nM, potentially leading to an overestimation of K_cat_/K_M_. This very efficient *trans-*cleavage rate supports its use for rapid, amplification-free detection.

### Multiple crRNAs in droplets report the presence of multiple target sequences

We next tested the effect of crRNA combinations, which can generate more signal per target RNA in bulk^1^, in the droplet format. We loaded *in vitro* transcribed (IVT) target RNA corresponding to the N gene of SARS-CoV-2 into droplets containing either crRNA 2, crRNA 4, which targets a different region of the N gene, or both crRNAs (Fig. 1F), and then quantified the number and signal of the positive droplets. Surprisingly, while crRNAs 2 and 4 generate similar signals when used individually and might be expected to double the signal when combined^1^, we found that the signal per droplet was not significantly different in any of the positive droplets (Fig. 1G). Rather, the number of positive droplets increased when droplets contain both crRNA 2 and crRNA 4 compared to just one of the crRNAs (Fig. 1H). This suggests that RNA corresponding to the IVT N-gene was fragmented by Cas13a cleavage upon initiation of the reaction prior to droplet formation, causing the regions targeted by different crRNAs to be loaded into separate droplets (Fig. 1F). The resulting fragmented loading we observe is consistent with a recent study of microwell-based Cas13a reaction^3^ and suggests that multiple crRNAs can activate independent single-Cas13a reaction in droplets.

### crRNA/target RNA combinations govern Cas13a reaction kinetics

The Cas13a kinetics observed in droplets critically depend on the crRNA and its target. For example, two other crRNAs targeting the SARS-CoV-2 N gene—crRNA 11 and crRNA 12— exhibit significantly slower rates in bulk reactions than crRNA 2 or 4 (Fig. 2A). For this reason, the selection of “good” crRNAs that maximize Cas13 activity is critical for bulk Cas13-based molecular diagnostics^17^, though how different crRNAs affect the activity of Cas13 is not well understood^18^. We therefore tested crRNAs 11 and 12 in our droplet assay and compared it to the activity of crRNA 4. After the droplet reaction was incubated for 15 min, we found that both the number of positive droplets and signal-per-droplet was reduced for crRNAs 11 and 12 compared to crRNA 4 (Extended Data Fig. 2A and 2B). By examining the signal time trajectories, we found that the three crRNAs and target sequences resulted in distinct kinetic behaviors, with the slope, shape, and x-intercept of the individual trajectories varying widely (Fig. 2B-E). In some trajectories, we found a strikingly stochastic behavior, with periods of no signal increase followed by periods of rapid signal increase (“rugged” vs smooth trajectory). Since each droplet contains, on average, a single target, these results indicate that specific crRNA/target combinations modulate Cas13a enzymatic activity at a single-molecule level.

**Figure 2:**
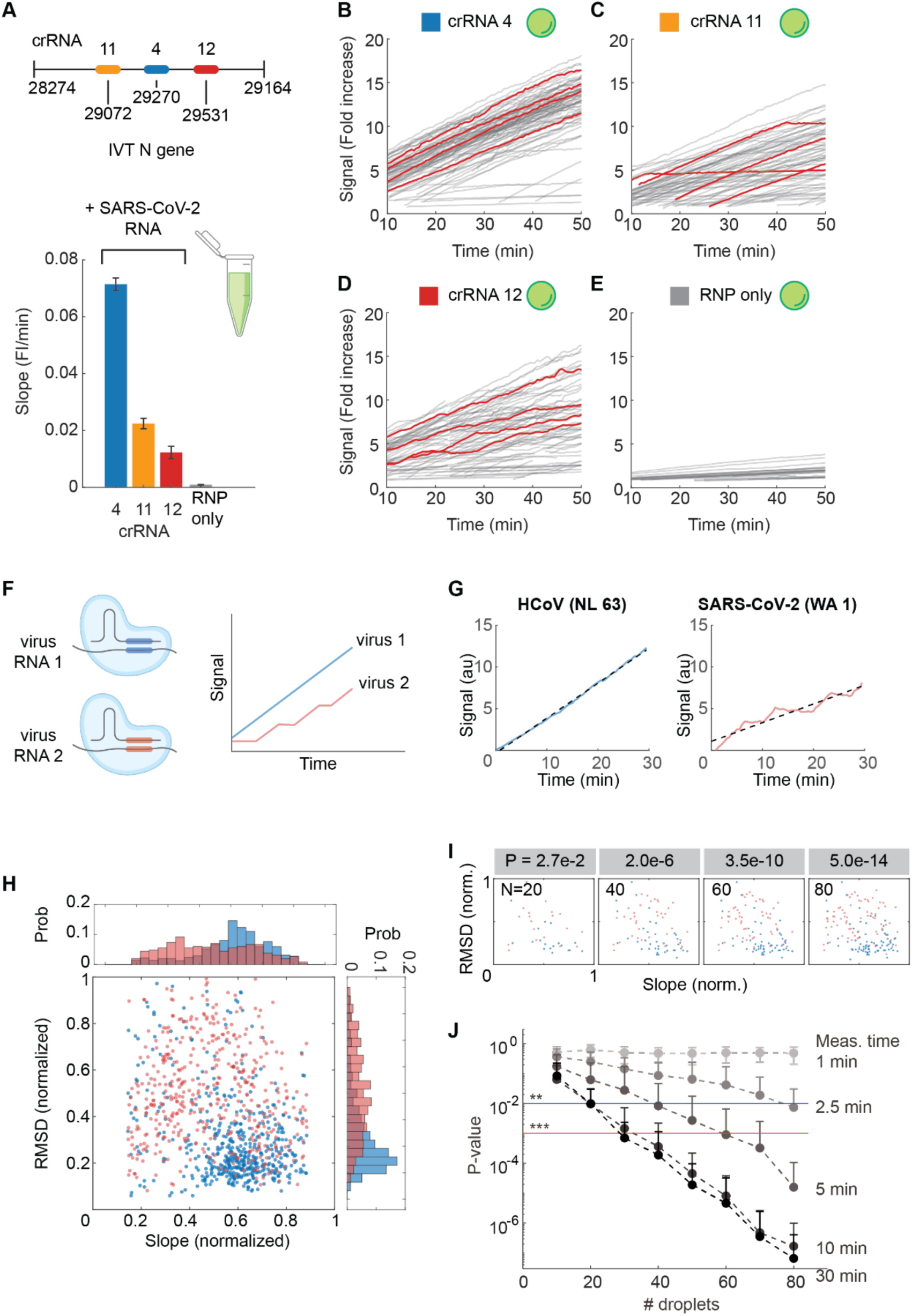
Cas13a kinetics are dependent on the crRNA and target RNA. a) The results of a bulk Cas13a reaction (fluorescence slope) with 3.5 x 10^4^ copies/µL of SARS-CoV-2 RNA for crRNAs targeting different regions of the SARS-CoV-2 N gene. The RNP only condition is measured with all three crRNAs. The slope is determined by performing simple linear regression of data from each replicate (N = 3) individually. Data are represented as mean ± SD. b) Time trajectories of single Cas13a reactions using crRNA 4. One hundred individual trajectories are measured in droplets ranging from 30 to 36 µm in diameter (grey lines), and representative trajectories are highlighted (red lines). The signal is measured every 30 seconds and represented as fold increase from the initial value. Two replicate experiments were combined for each crRNA. c) The same experiment as described in b), but with crRNA 11. d) The same experiment as described in b), but with crRNA 12. e) Time trajectories of droplets containing no target RNA but generating a non-zero signal from a Cas13a reaction. Thirty-one individual trajectories from two replicate runs were measured in droplets ranging from 30 to 36 µm in diameter. f) Schematic showing the principle of kinetic barcoding for detection of two different viruses in the same droplet reaction. It relies on the unique Cas13a kinetic signatures for specific combinations of a crRNA and target. g) Representative single Cas13a reaction trajectories with HCoV-NL-63 RNA (targeted by crRNA 7) or SARS-CoV-2 RNA (targeted by crRNA 12). The signal is the total fluorescence change in a droplet normalized by its initial value, which remains invariant regardless of droplet size. The dotted black line shows a linear fit. h) Distribution of slope and RMSD of individual Cas13a trajectories between HCoV-NL-63 and SARS-CoV-2 RNA. The slope and RMSD is determined from individual 30-minute-long trajectories (N = 551 for HCoV-NL-63 and N = 488 for SARS-CoV-2) measured with a single crRNA-target pair. The normalized RMSD is rescaled from 0 to the highest value of the dataset. The slope is rescaled from 0 to the highest value in the dataset (blue = HCoV-NL-63; red = SARS-CoV-2). i) Identification of HCoV-NL-63 or SARS-CoV-2 based on the kinetic parameters of individual Cas13a reactions. Varying numbers of 30-minute-long Cas13a trajectories are randomly selected from each condition, and the difference between two groups are quantified as p-values based on a two-tailed *Student’s* t-test. j) The accuracy of kinetic barcoding quantified as p-values based on a two-tailed *Student’s* t-test. P-values are determined for decreasing measurement times by taking the initial parts of Cas13a trajectories. Data shows mean ± SD of 50 repeated iterations. ** (blue line) p < 0.005, *** (red line) p < 0.001.

To quantify these kinetic differences, we characterized individual trajectories by their average slope, root-mean-square-deviation (RMSD), and time from target addition to initiation of enzyme activity (T_init_) (Extended Data Fig. 2C). By calculating the instantaneous slopes at each point in the trajectory and fitting their distribution to a Gaussian, we found that the trajectories exhibited two different slopes, one “fast” when fluorescence was increasing and one “slow” when fluorescence was not increasing (Extended Data Fig. 2D). Interestingly, even though the average slopes differed significantly for the different crRNAs, the instantaneous “fast” slopes were constant across all three crRNAs (Extended Data Fig. 2E). Consistent with this, crRNA 12, which exhibited the lowest average slope of the three crRNAs, showed extended “slow” periods (Extended Data Fig. 2F) and increased level of signal fluctuation (Extended Data Fig. 2G). We found that T_init_ varied significantly among the three crRNAs we tested, with crRNA 11 exhibiting the slowest T_init_ (Extended Data Fig. 2H), resulting in reduced signal at the reaction endpoint (Extended Data Fig. 2B). To test if the stochastic behavior of individual Cas13a reactions is simply caused by the unbinding of crRNAs from the Cas13a protein, we changed the ribonucleoprotein (RNP) concentration to below (for crRNA4) or above (for crRNA12) the K_d_ of the crRNA-Cas13a complex^9,19^ but the stochastic behavior was unaltered (Extended Data Fig. 2I and 2J). In contrast, when we changed the target from genomic SARS-CoV-2 RNA to a 20-nucleotide fragment complementary to crRNA 12’s spacer sequence, the stochastic behavior of the reaction was no longer observed, and T_init_ was significantly shortened (Extended Data Fig. 2K). This suggests that the kinetic signature of a specific crRNA/target RNA is robust regardless of the variations in the reaction conditions but is sensitive to the crRNA/target RNA sequence and the target RNA length.

### Heterogeneous Cas13a kinetics offer a method for multiplexed detection

We hypothesized that the distinct kinetic signatures of the different crRNAs could be used to identify different crRNA-target pairs within a droplet, thus providing a method for multiplexed detection of different RNA viruses (Fig. 2F). To test this idea, which we termed ‘kinetic barcoding’, we combined a crRNA targeting a common cold virus HCoV-NL-63 (HCoV crRNA 6) and a second crRNA targeting SARS-CoV-2 (crRNA 12). The crRNAs were chosen because they individually exhibit different kinetic signatures on their respective targets (Fig. 2G). We collected 30-minute trajectories from hundreds of droplets containing either HCoV-NL-63 or SARS-CoV-2 viral RNA along with Cas13a and both crRNAs together. The two groups of trajectories associated with the two different crRNAs were clearly distinguishable based on their average slope and RMSD (Fig. 2H). On an individual basis, trajectories for HCoV-NL-63 and SARS-CoV-2 were discerned with a 75% accuracy using a support vector machine (SVM) for binary classification (Methods). To determine how well two viruses can be distinguished when multiple trajectories are combined, we randomly sampled a subset of trajectories and compared their difference by performing Student’s t-test on their binary classification result (Fig. 2I, Methods). This analysis showed that kinetic barcoding can distinguish between HCoV-NL-63 and SARS-CoV-2 targets within 10 minutes provided that 20 or more trajectories (i.e. 20 or more positive droplets) are measured (Fig. 2J).

### DNA modifications of crRNA can programmably tune Cas13a kinetics

The experiments above with naturally varying enzyme kinetics show the potential of kinetic barcoding for multiplexed detection. However, searching for crRNA/target pairs that exhibit distinct kinetic signatures could be time consuming and is not guaranteed, especially in the case of variants with small numbers of mutations. Building on the observation that regions outside the spacer region affect Cas13a kinetic activity^20^, we hypothesized that positioning a single-stranded DNA fragment proximal to Cas13a’s HEPN domain will interfere with its *trans*-cleavage activity for RNA without being digested, thus slowing down the rate of reporter RNA cleavage. To test this, we added a DNA fragment of varying length and sequence to the 5’-end of crRNA 4, which was sufficiently long that it could reach the HEPN domain when the crRNA is bound to Cas13a (Fig. 3A). We referred to this hybrid RNA-DNA guide as an ‘interfering gRNA’, or igRNA. We empirically divided the igRNA into an effector region, which can alter Cas13a’s nuclease activity, and an 8bp-long linker region that connects the effector DNA and crRNA. We measured the reporter signal in a solution containing either the igRNA 4 or unmodified crRNA 4 along with synthetic SARS-CoV-2 RNA targets and found that Cas13’s *trans*-cleavage rate was reduced for igRNA 4, with DNA effectors having longer lengths and thymines (T) showing stronger reductions (Fig. 3B). The same reduction in signal slope was also observed in single droplets, confirming the change in Cas13 *trans*-cleavage activity at a single-molecule level (Extended Data Fig. 3A). Our result is consistent with the prediction that increasing the local concentration of DNA near the HEPN domain (by adding more nucleotides at the 5’-end of crRNA) or addition of DNA sequences matching the *trans*-cleavage preference (by including thymines in the effector region) will interfere with its nuclease activity more strongly. We concluded that our crRNA modification strategy, which can be used with any target sequences, enables precise tuning of Cas13a kinetics at a single-molecule level.

**Figure 3:**
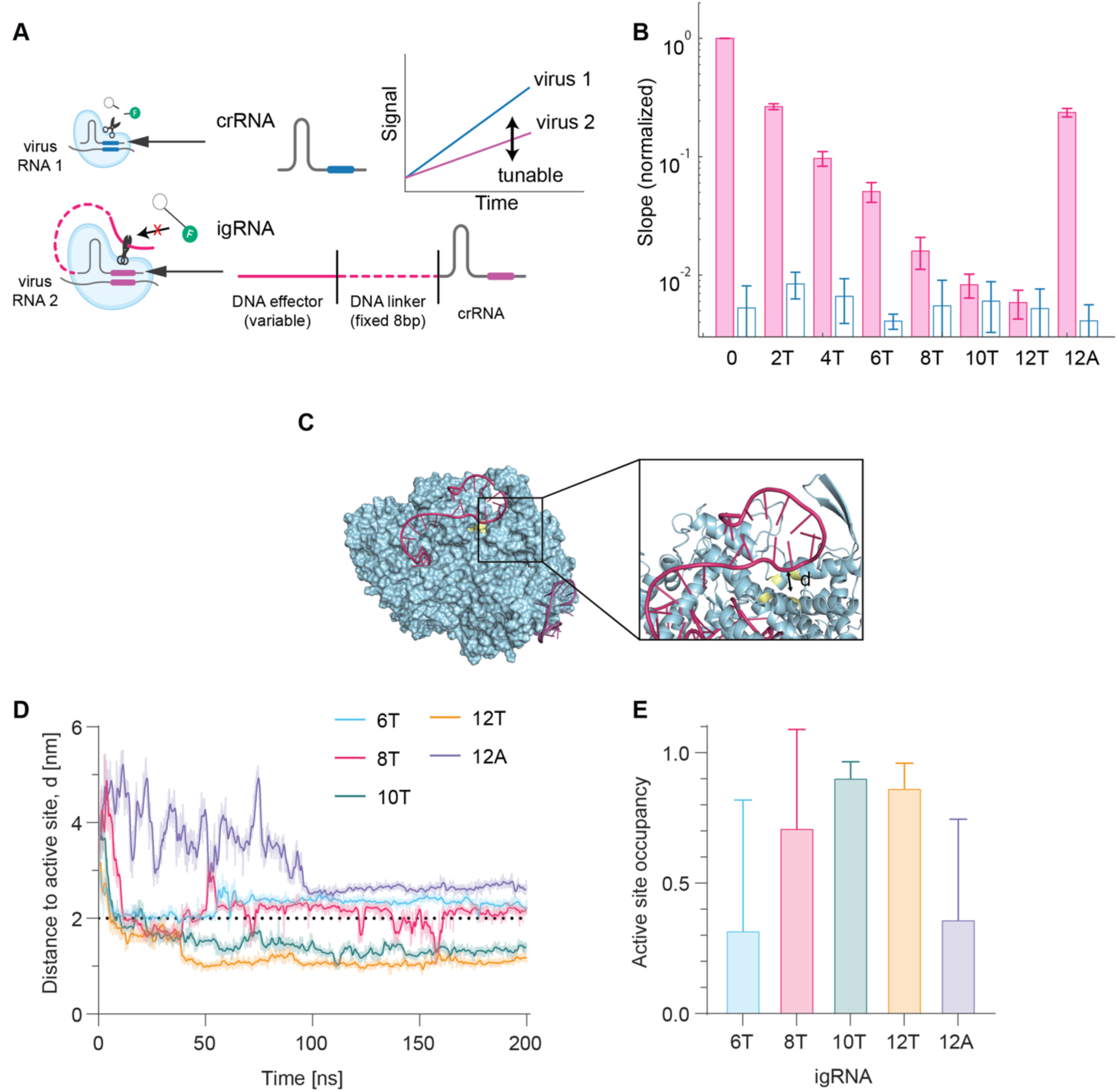
Kinetic barcoding for multiplexed detection of viruses and variants. a) Schematic showing the principle of kinetic barcoding with igRNA. DNA effector of varying sequence and length is added in the 5’-end of crRNA through a fixed, 8bp-long DNA linker, leading to interference with the HEPN nuclease activity. b) Bulk Cas13 reaction rate is altered by igRNA. DNA effector with varying number of Ts or 12A are added to crRNA4 and the reaction rate is measured with a fixed amount of synthetic target RNA (filled bar) or RNP only (hollow bar). The slope is normalized by the slope measured with unmodified crRNA4 and presented in logarithmic scale. Data are represented as mean ± SD of three replicates. c) DNA effector blocks LbuCas13a active site. Representative snapshot at t=200 ns of molecular dynamics simulation for 12T igRNA. The igRNA (red) residues position themselves directly on top of the LbuCas13a (light blue) active site (yellow). Inset: d represents the shortest distance from DNA effector of igRNA to the center of mass for the catalytic site (R472, H477, R1048, H1053). d) Dynamics of active site occupancy by DNA effector represented as d over time. Solid lines represent moving average over 1 ns simulation time and translucent lines represent raw data. e) Occupancy of active site changes based on DNA effector length and sequence. Active site occupancy as a fraction of total simulation time (200 ns) for each igRNA. Error bars correspond to standard deviation from 3 independent replicate runs.

To investigate the mechanism of this inhibitory behavior, we conducted atomistic molecular dynamic simulations of the 6T, 8T, 10T, 12T, and 12A igRNAs (See Methods). We found for all igRNAs, the 5’ end of the DNA effector moved towards the active site, effectively blocking it (Fig. 3C, 3D). For all igRNAs, the effector region approached the active site very rapidly. However, we noticed thymine-based igRNAs converged faster (∼25 ns) compared to the 12A igRNA (∼100 ns) (Fig. 3D). This observation suggests there are mostly sequence agnostic interactions between the linker region and LbuCas13a which bias movement of the DNA effector towards the active site. Furthermore, we noticed the 8T, 10T, and 12T igRNAs all reached distances significantly closer than the 6T and 12A igRNA supporting previous observations regarding length and sequence constraints needed for effective suppression (Fig. 3B).

We then sought to determine the residence time for each DNA effector near the active site. We classified the active site as occupied when any nucleotide from the DNA effector was within a 2.0 nm radius of the active site (See Methods for distance calculations). We found that in agreement with our experimental results (Fig. 3B), active site occupancy increased proportionally with DNA effector length for thymine-based igRNAs (Fig. 3E). The 8T, 10T, and 12T igRNAs showed more consistent active site occupancy than the 6T igRNA or 12A igRNA (Fig. 3E), suggesting sequence selectivity occurs in the surface region near the active site (Supplementary Videos 2, 3, and 4). Although we noticed similar occupancies between the 8T, 10T, and 12T igRNAs, we reason at longer timescales, the larger amount of thymines able to reach the active site for the 12T igRNA explains the higher suppression for this igRNA compared to the 8T and 10T. We speculate the mechanism for this sequence preference reflects LbuCas13a’s previously characterized nucleotide cleavage preferences^20^.

We next tested whether our programmable kinetic barcoding strategy can improve multiplexed virus detection. We first selected 4 different crRNAs targeting different virus RNA that all provided identical *trans*-cleavage rates for their respective target (crRNA 4 for SARS-CoV-2 wildtype, crRNA delta for SARS-CoV-2 delta, crRNA NL63 for HCoV-NL-63, and crRNA H3N2 for H3N2 influenza virus), and hence could not be used to distinguish viral targets based on kinetics in a droplet Cas13 assay (Extended Data Fig. 3B). We then added effector DNA of increasing lengths to each crRNA and measured 1-hour trajectories from droplets containing individual target virus RNA. We observed that the trajectories were linear, and their slopes were clearly separated from one virus to another (Fig. 4A, Extended Data Fig. 3C). Importantly, the slopes for each individual virus remained the same even when all four crRNAs were combined into the same droplet (Extended Data Fig. 3D and 3E), making it possible to simultaneously detect different targets based on their unique slopes. One exception was SARS-CoV-2 delta, which showed two peaks when the crRNAs were combined since both crRNA 4 and crRNA delta targeted its RNA. We focused on the three viruses exhibiting a single peak with the crRNA-combination and classified samples containing either one or two different viruses based on their slopes (see Methods). We found that our programmable kinetic barcoding approach not only correctly identified the target but also quantified the proportion of each target among the possible single or dual infection scenarios (Fig. 4B, Extended Data Fig. 3F).

**Figure 4:**
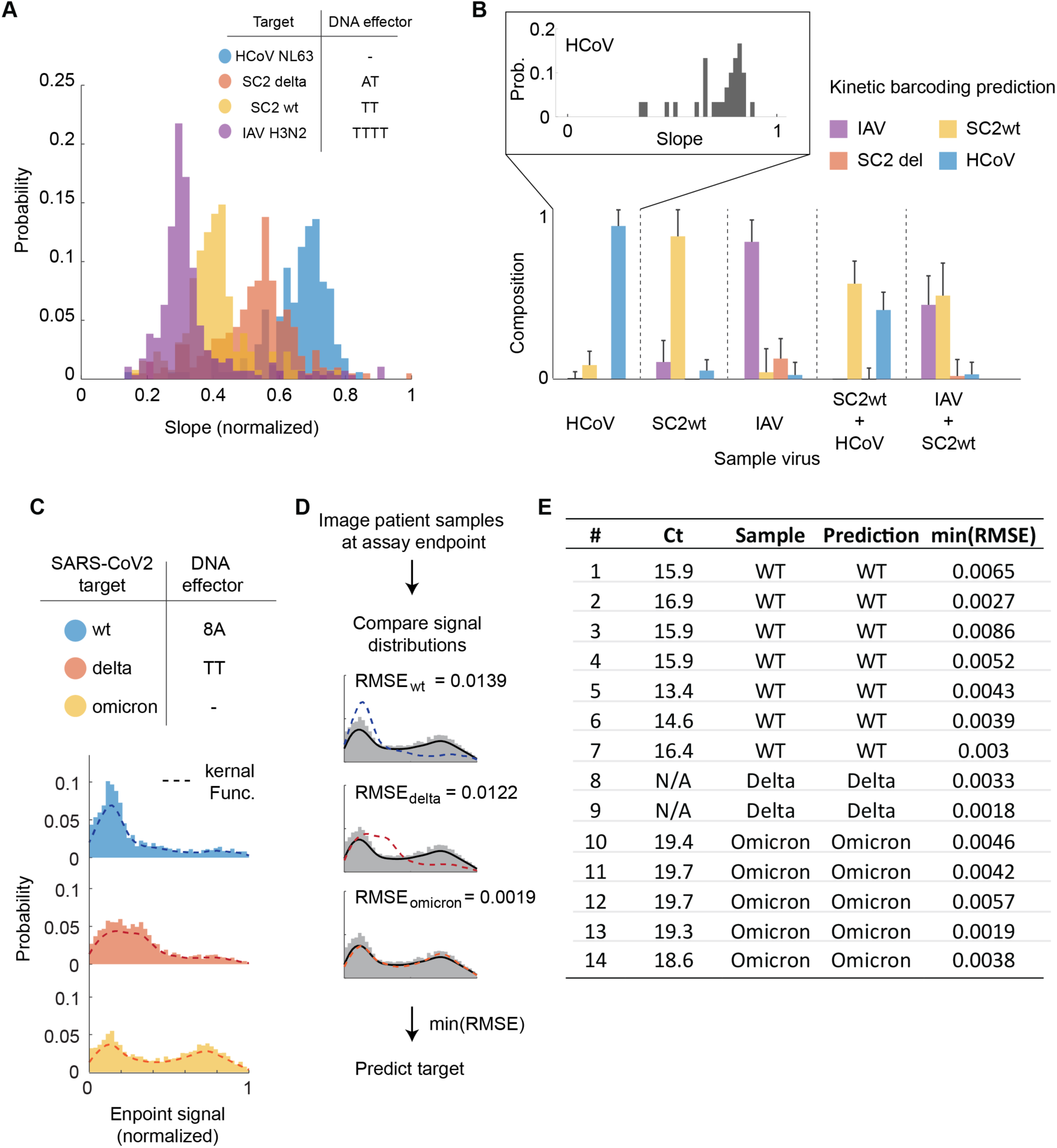
Endpoint kinetic barcoding with clinical samples. a) Measurement of 4 different viruses with modified crRNAs containing different DNA effector (Top). Total N from two replicate measurements were 169, 406, 128, and 285 for HCoV NL63, SARS-CoV-2 (SC2) delta, SC2 wt, and IAV H3N2. Signal slope is normalized for droplet size and its distribution is shown in histogram. b) Prediction of target viruses based on the signal slope distribution. From more than 100 trajectories obtained for each sample in two replicate experiments, 30 trajectories are randomly selected to mimic samples with intermediate virus concentrations (representative distribution shown in the inset and in Extended data Fig. 3F) and the virus compositions in the sample are predicted. The bar graph shows the mean and standard distribution of target predictions from 100 repeated sampling. c) Design of crRNAs for multiplexed SC2 variant detection (top). Droplet Cas13a reaction is incubated with an individual variant’s synthetic RNA to obtain the standard curve (bottom). The dotted line indicates the kernel function of slope distribution. d) Clinical sample analysis scheme. The kernel function of clinical samples is compared with the standard curves obtained in a). The one that provides the smallest difference, measured in terms of RMSE between two curves, is selected as the target. e) Summary of clinical samples and kinetic barcoding prediction result. The Ct-values are obtained from the N-gene. For SC2 delta virus, RNA isolated from a cell was used (BEI resources) instead of patient sample.

### Endpoint kinetic barcoding can accurately differentiate virus variants

From a practical perspective, evaluating a multiplexed RNA detection assay with a single measurement would be much preferred over the requirement to collect a series of measurements over time. To that end, we explored whether kinetic barcoding was a viable strategy if only the endpoint fluorescence of droplets were measured rather than their full-time trajectory, a realistic possibility since the programmable kinetic barcoding strategy alters only reaction slope without introducing additional stochasticity. We first built a rapid imaging platform that consisted of a low magnification objective (2.5X / NA 0.085) and an automated stage that could image the approximately 160,000 droplets from a single 20µl reaction in 45 seconds (see Methods). We then tested whether endpoint kinetic barcoding could be used with patient samples to differentiate two dominant SARS-CoV-2 variants circulating between November 2020 and January 2022 – delta and omicron – from wildtype SARS-CoV-2. In addition to crRNA 4 targeting a conserved region of SARS-CoV-2 RNA, we chose two crRNAs that are specific to unique mutations in delta (crRNA delta) or omicron (crRNA omicron) and added effector DNA modifications to differentiate their kinetics (Fig. 4C). We tested different virus targets with the 3-crRNA combination and imaged the droplets after one-hour incubation using the endpoint detection system. As expected, the wildtype virus RNA showed a single peak distribution while the delta or omicron RNA exhibited two peaks – one corresponding to the variant and the other to the shared region of SARS-CoV-2 genome (Fig. 4C). To classify each clinical sample, we compared its signal distribution to the three reference curves we collected and identified the one that exhibited the least difference as determined by RMSE or the Kolmogorov–Smirnov test (Fig. 4D, Extended Fig. 4A). Focusing on samples with mid-to-high virus concentrations (Ct < 20), we tested 15 positive SARS-CoV-2 samples and accurately classified all samples through kinetic barcoding (Fig. 4E, Extended Data Fig. 4B).

## Conclusion

In summary, we demonstrate that a droplet-based Cas13a direct detection assay can distinguish RNA targets based on single-Cas13 reaction kinetics. We found that LbuCas13a is an efficient, diffusion-limited enzyme, whose kinetics are controlled by the specific combination of the crRNA and the target^21,22^. Interestingly, the distribution of Cas13a RNP trajectories is largely homogenous for crRNAs supporting high enzymatic activities (Fig. 2B), implying that the active conformation of Cas13a RNP is stable over time. However, certain crRNAs were found to induce stochastic trajectories on the timescale of minutes. The observation that this kinetic feature is abolished when a short RNA target is used instead of the longer viral RNA (Extended Data Fig. 2K) suggests a role of local or global RNA structure on modifications in the kinetics of an individual Cas13a.

We first took advantage of these characteristic kinetic signatures to show that different viral targets could be distinguished in a single droplet. We then devised a simple crRNA modification strategy that enables programmable tuning of LbuCas13’s *trans*-cleavage rate, expanding the diagnostic utility of kinetic barcoding and the number of targets that could be distinguished simultaneously. Kinetic barcoding relies on reading the crRNA/target RNA-dependent single-Cas13a activity, which was possible because the early fragmentation of a target RNA before droplet formation generated multiple droplets containing a single, distinct target fragment (Fig. 1F). We demonstrate this principle by multiplexing four targets, but we anticipate kinetic barcoding of more can be achieved with further development of the assay and LbuCas13a itself, as well as by combining natural and programmed kinetic barcoding.

The kinetic barcoding strategy we present represents a new approach to multiplexing based on single enzyme kinetics, adding a new dimension to RNA detection that could enhance existing multiplexing strategies. Previous CRISPR-Cas-based assays achieved multiplexed detection by employing multiple Cas orthologs acting on different color reporters^10^ or massively dividing reactions after amplification into droplets that each detect only a single target^23^. Kinetic barcoding could be readily combined with color-based methods to immediately achieve multiplexing of more than 10 target pathogens or with the multiplexed droplets to act as a multiplier. A deeper understanding of the kinetics and mechanisms of Cas13 and other CRISPR nucleases will also offer new opportunities in diagnostic and therapeutic development. .

## DATA AVAILABILITY

The data that support the findings of this study are available from the corresponding author upon reasonable request.

## Data Availability

All data produced in the present study are available upon reasonable request to the authors

## ACKNOWLEDGEMENTS

We thank all members of the Fletcher, Ott, and Doudna laboratories for helpful discussions and feedback on this project. We also thank Stacia Wyman, Erica Moehle, and Bryan Bach for providing samples from the Innovative Genomics Institute, and Chaz Langelier for providing samples from the Chan-Zuckerberg Biohub. Purified LbuCas13a was a kind gift from Shanghai ChemPartner, and we thank Synthego for support with synthetic crRNAs. We gratefully acknowledge support from NIH/NIAID grant 5R61AI140465-03 to J.A.D., D.A.F., and M.O. and a generous gift from an anonymous donor in support of the ANCeR diagnostics consortium, as well as generous individual donors to the Gladstone Institutes. This work was supported in part by the Health Tech CoLab at the Blum Center for Developing Economies at UC Berkeley. G.J.K was supported by the Snow Medical Research Foundation (SMRF2021-276) and an NHMRC Investigator Grant (EL1, APP1175568). J.A.D. is an HHMI Investigator. D.A.F. and M.O. are Chan-Zuckerberg Biohub Investigators. The following reagents were deposited by the Centers for Disease Control and Prevention and obtained through BEI Resources, NIAID, NIH: Genomic RNA from SARS-Related Coronavirus 2, Isolate USA-WA1/2020, NR-52285; SARS-Related Coronavirus 2, Isolate USA-WA1/2020, NR-52281; SARS-Related Coronavirus 2, Isolate hCoV-19/USA/MD-HP05285/2021 (Lineage B.1.617.2; Delta Variant), NR-56127; and Human Coronavirus, NL63, NR-470.

## AUTHOR CONTRIBUTIONS

S.S., D.A.F., and M.O. conceived and designed the study. S.S., A.L., and C.F.N. performed experiments and S.S. analyzed data. J.S., S.I.S., P.F., G.R.K., G.J.K. designed crRNAs. D.B. and C.S. assisted with BSL-3 work. J.S., S.I.S., P.F., G.R.K., G.J.K., D.C.J.S., T.Y.L. and J.A.D provided reagents. G.R.K., J.A.D., and M.O. obtained clinical samples. G.J.K., T.Y.L., and J.A.D. provided critical expertise and feedback. A.D. performed molecular dynamics simulations. D.A.F. and M.O. supervised the study design and data collection. J.A.D., M.O., and D.A.F. secured funding. S.S. and D.A.F. wrote the manuscript, and all authors contributed feedback.

