## Supplemental Materials for "Programmable kinetic barcoding for multiplexed RNA detection with Cas13a"

Son, et al.

This file contains Extended Data Figures 1-4, Supplementary Videos 1-3, and Supplementary Table 1.

EXTENDED DATA FIGURES

**Extended Data Figure 1**

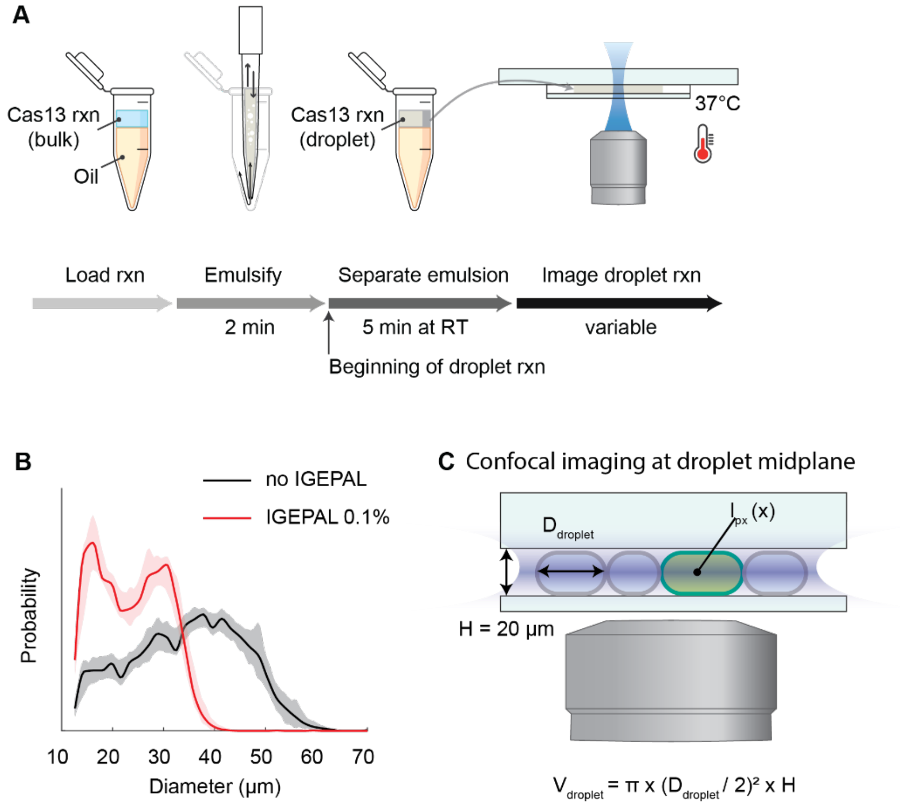

**Extended Data Fig. 1: Formation and imaging of Cas13 droplets**

a) Workflow of the droplet Cas13a assay. A Cas13a reaction including target RNAs is mixed with an oil (HFE 7500 including 2% w/w Perfluoro-PEG surfactant) and emulsified by repeated pipette mixing at a constant speed for 2 minutes. The emulsified reaction is held at room temperature for 5 minutes to wait for the emulsion to be separated from the aqueous phase. Subsequently, the emulsion is loaded into a custom imaging chamber and imaged with a fluorescence microscope.

b) Droplets containing the Cas13a reactions range in size from approximately 15 to 40 µm. The droplet size distribution (red line) is increased when the 0.1% wt/vol IGEPAL is removed from the Cas13a mix (black line). The shadows indicate the S.D. from 5 independent droplet preparations

c) Schematic depicting the geometry of flow cell and droplets, and confocal imaging of fluorescence signal (not drawn to scale). A 20X Water Immersion objective was used when droplet volume needed to be accurately determined. $D_{droplet}$ = droplet diameter, $V_{droplet}$ = droplet volume, $I_{px}(x)$ = fluorescence signal intensity of camera pixel in location x. Droplets larger than 20 µm deform to disks.

**Extended Data Figure 2**

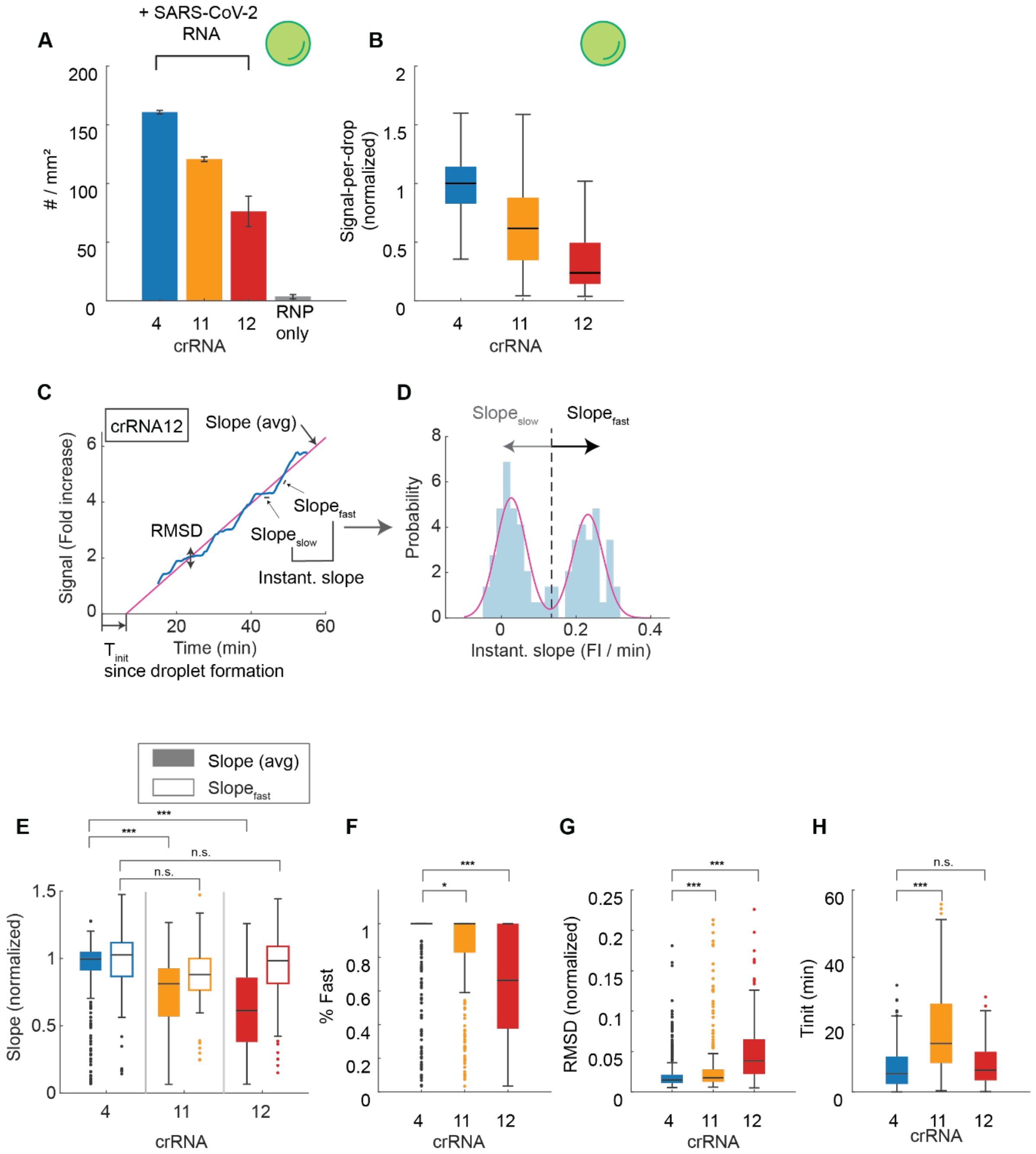

I

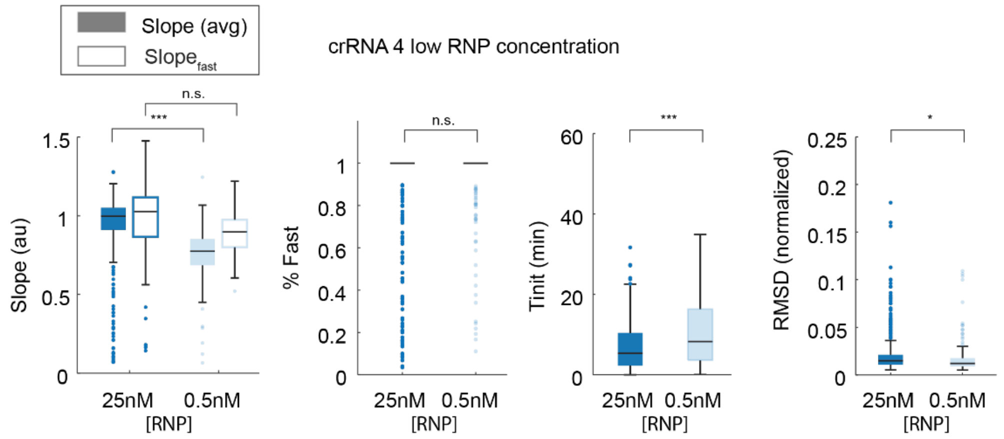

Slope (avg)
Slope fast
crRNA 4 low RNP concentration
n.s.
***
Slope (au)
% Fast
Tinit (min)
*
RMSD (normalized)
25nM 0.5nM
[RNP]


J

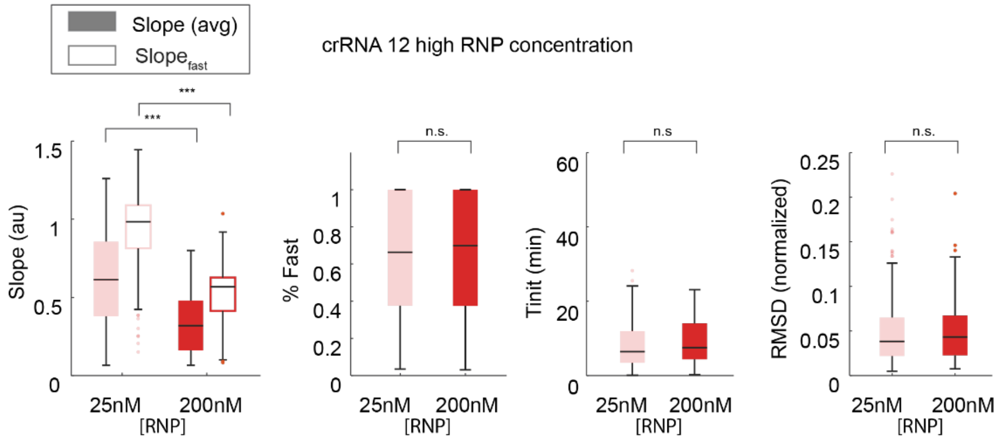

Slope (avg)
Slope fast
crRNA 12 high RNP concentration
***
Slope (au)
n.s.
% Fast
n.s
Tinit (min)
RMSD (normalized)
25nM 200nM
[RNP]


K

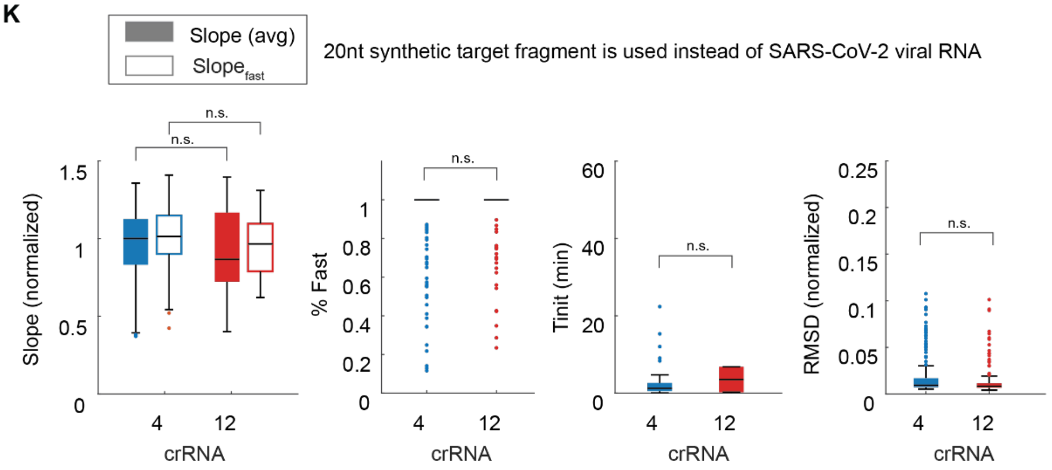

Slope (avg)
Slope fast
20nt synthetic target fragment is used instead of SARS-CoV-2 viral RNA
n.s.
Slope (normalized)
% Fast
Tinit (min)
RMSD (normalized)
4 12
crRNA

**Extended Data Fig. 2: crRNA/target RNA-dependent Cas13a kinetics.**

a) The results of droplet Cas13a reactions (# positive droplets / $mm^2$) with 3.5 x $10^4$ copies/µL of SARS-CoV-2 RNA for crRNAs 4, 11, and 12 after 30 minutes of incubation. The RNP only condition is measured with all three crRNAs. A total of 1287, 966, 605, and 21 positive droplets were quantified from each reaction. Data are represented as mean ± SD of three replicates except for crRNA 4, where two replicate experiments were performed.

b) The same experiment as described in a) but reporting the average fluorescence signal in each droplet containing the different crRNAs after 30 minutes. The signal-per-droplet is represented as a box and whisker plot showing the median, lower and upper quartiles, and minimum and the maximum values ignoring outliers. The signal is normalized by the median of crRNA 4.

c) Analysis strategy of an individual Cas13a signal trajectory, using crRNA 12 as an example trajectory (blue curve). The average slope (Slope (avg)) (red curve), $T_{init}$, and the Root-mean-square-deviation (RMSD) are determined by performing simple linear regression to the raw signal. $Slope_{fast}$ and $Slope_{slow}$ correspond to the fast and slow periods determined in d).

d) Instantaneous slopes of the Cas13a signal trajectories are calculated by taking the time-derivative of the raw signal (blue histogram) and fitting its probability distribution with either single- or binary-Gaussian (red line). For data that favors binary distribution, we determined $Slope_{fast}$, $Slope_{slow}$, % Fast, and % Slow by taking the mean and the proportion of each Gaussian.

e) The distribution of slopes is represented as a box and whisker plot showing the median, lower and upper quartiles, and the minimum and the maximum values, with outliers overlaid as individual data points. Individual 30-minute-long trajectories from droplets of arbitrary diameter are used after their signal is normalized for droplet size. To calculate the distributions, 395, 267, and 311 trajectories are used for crRNA 4, crRNA 11, and crRNA 12, respectively. P-values determined from a two-tailed *Student's* t-test are 2.1e-20, 7.1e-45, 0.34, 0.39. ns = not significant, *$p < 0.05$, ***$p < 0.001$.

f) The same experiment as described in e), but for % fast. P-values determined from a two-tailed *Student's* t-test are 0.021 and 4.3e-34.

g) The same experiment as described in e), but for RMSD. P-values determined from a two-tailed *Student's* t-test are 1.5e-4 and 4.4e-30.

h) The same experiment as described in e), but for $T_{init}$. P-values determined from a two-tailed *Student's* t-test are 1.6e-23 and 7.7e-2Bulk Cas13a reaction rate for 4 x $10^5$ copies/µL of *in-vitro* transcribed (IVT) N gene RNA was measured with crRNA 2 or 4 and represented as mean ± SD of three replicates.

i) Effect of RNP concentration on Cas13a reaction kinetics using crRNA 4. Distribution of kinetic parameters is represented as box and whisker plots representing the median, the lower and upper quartiles, and the minimum and the maximum values, with outliers overlaid as individual data points. 395 and 190 individual 30-minutes-long trajectories are collected from two replicate experiments for each RNP concentrations. P-values determined from a two-tailed *Student's* t-test are 8.8e-18 and 0.13 (slope), 0.37 (% Fast), 9.2e-5 ($T_{init}$), and 1.1e-2 (RMSD). ns = not significant, *p < 0.05, ** p < 0.005, ***p < 0.001.

j) As in i), but for crRNA 12. We collected 311 and 102 individual trajectories from two replicate experiments for each RNP concentration. P-values determined from a two-tailed *Student's* t-test are 1.4e-17 and 6.1e-29 (slope), 0.69 (% Fast), 0.39 ($T_{init}$), and 0.81 (RMSD). ns = not significant, *p < 0.05, ** p < 0.005, ***p < 0.001.

k) Effect of target RNA size on Cas13a reaction kinetics. Distribution of key parameters is represented as the box and whisker plots representing the median, the lower and upper quartiles, and the minimum and maximum values, with outliers overlaid as individual data points. We collected 230 and 129 individual 30-minutes-long trajectories from two replicate experiments for each crRNA. P-values determined from a two-tailed *Student's* t-test are 0.35 and 0.34 (slope), 0.61 (% Fast), 0.21 (RMSD), and 0.91 ($T_{init}$). ns = not significant.

**Extended Data Figure 3**

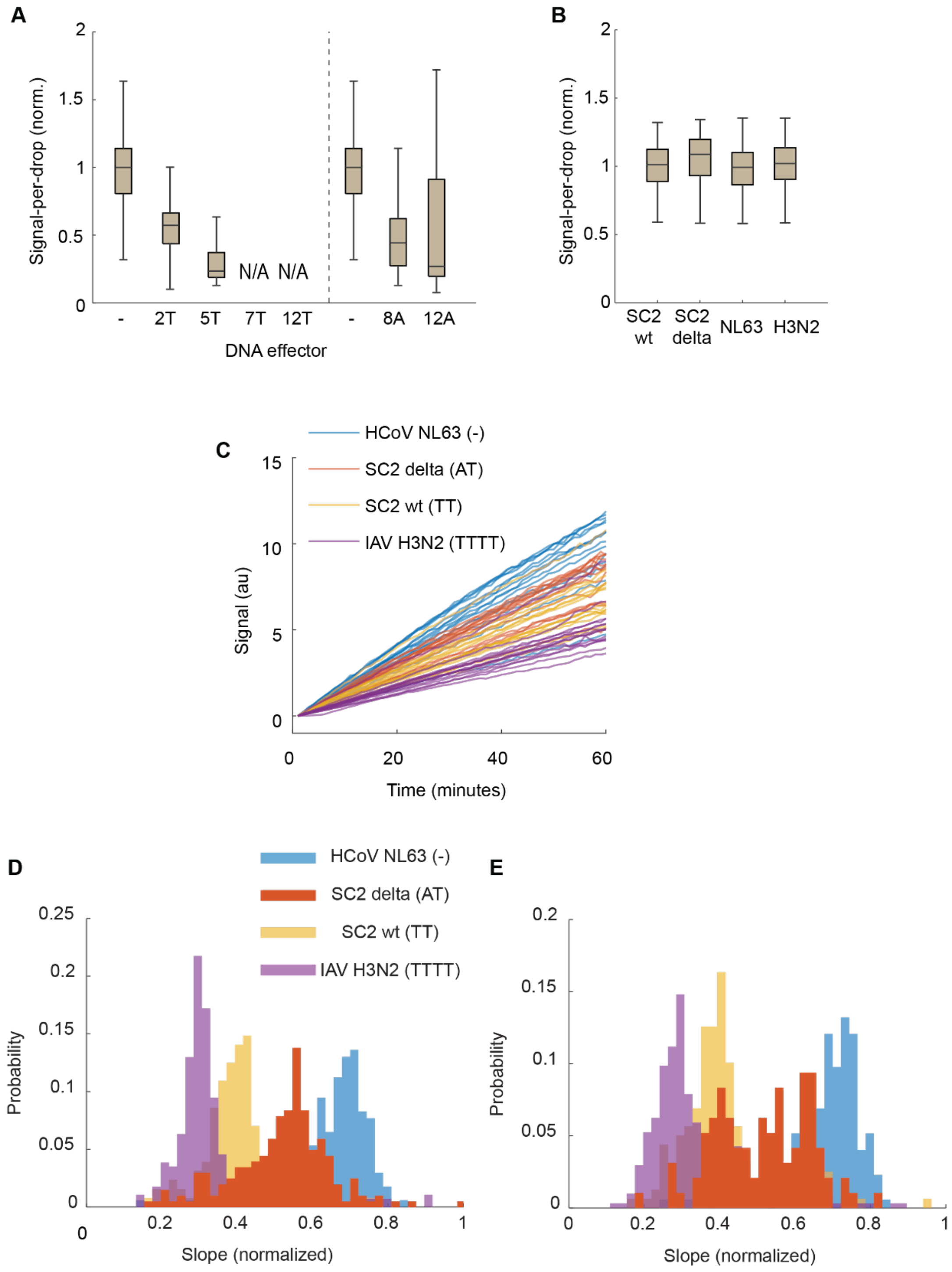

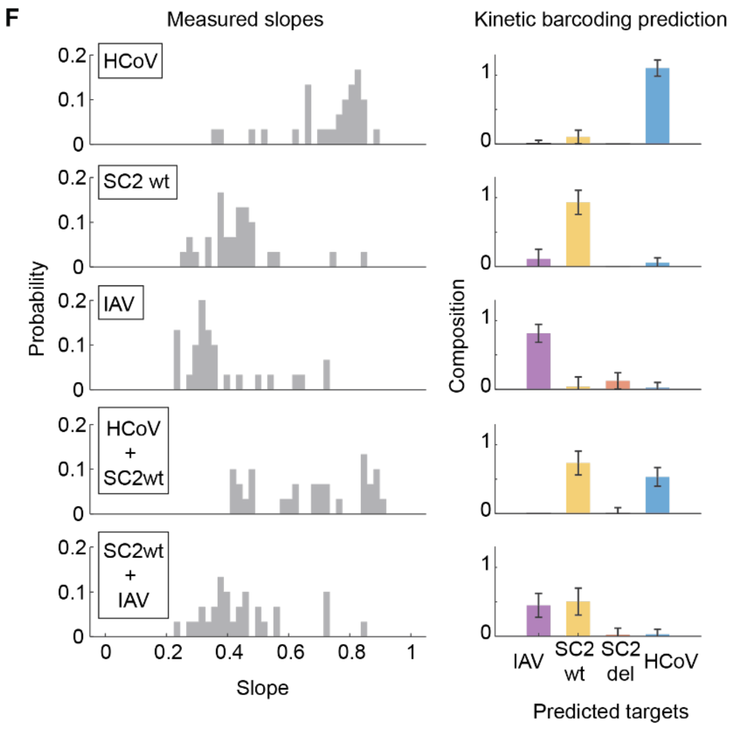


**Extended Data Fig. 3: crRNA modifications alter single-Cas13a activity.**

a) Effect of DNA effector on the droplet signal represented as a box and whisker plot showing the median, lower and upper quartiles, and the minimum and maximum values ignoring outliers. All DNA modifications are made to the same crRNA (crRNA4) and, for each crRNA design, droplet signal from SARS-CoV-2 RNA is quantified from individual droplets at the assay endpoint. N=1525 (-), 1114 (2T), 942 (5T), 1368 (8A), 484 (12A). No positive droplets are detected for crRNAs modified with 7T and 12T.

b) Droplet signal of different viruses measured with unmodified crRNAs. N=263 (SARS-CoV-2 (SC2) wt), 226 (SC2 delta), 1119 (NL63), and 553 (H3N2).

c) Raw signal time-trajectories are shown for four DNA-modified crRNAs targeting HCoV NL63, SC2 wt, SC2 delta, and IAV H3N2. The signal is normalized for droplet size and its initial value is fixed as 0. 15 representative trajectories are shown for each virus.

d) Histogram showing the signal slope distribution of an individual virus measured with the single crRNA.

e) Histogram showing the signal slope distribution of an individual virus measured with 4-crRNA combinations.

f) Prediction of target viruses based on the signal slope distribution. Total 268, 272, 518, 157, 119 trajectories were obtained for HCoV, SC2 wt, IAV, HCoV+SC2wt, and SC2wt+IAV, respectively. Among those, 30 trajectories are randomly selected from each sample (representative distribution shown on the left) and the target virus compositions are predicted from the subset. The bar graph on the right shows the mean and standard distribution of target predictions from 100 repeated sampling.

**Extended Data Figure 4**

A

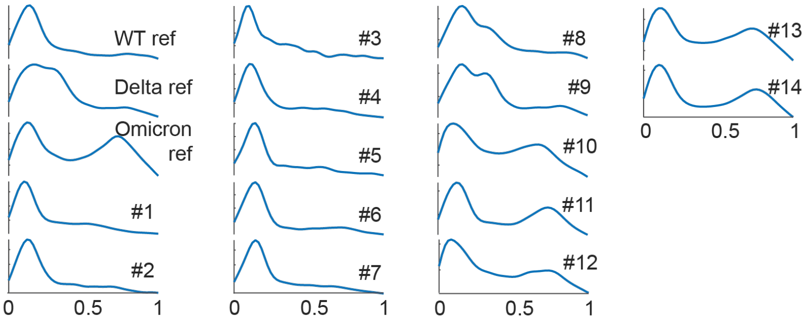


B

| Sample # | Ct | Sample | Difference (Kernel) | | | Difference (K-S) | | | Prediction |
|---|---|---|---|---|---|---|---|---|---|
| | | | WT | Delta | Omicron | WT | Delta | Omicron | |
| WT ref | N/A | WT | 0.0000 | 0.0115 | 0.0148 | 1.00E+00 | 1.29E-37 | 9.75E-76 | WT |
| Delta ref | N/A | Delta | 0.0115 | 0.0000 | 0.0130 | 1.29E-37 | 1.00E+00 | 5.15E-93 | Delta |
| Omicron ref | N/A | Omicron | 0.0148 | 0.0130 | 0.0000 | 9.75E-76 | 5.15E-93 | 1.00E+00 | Omicron |
| 1 | 15.9 | WT | 0.0065 | 0.0138 | 0.0157 | 3.51E-10 | 2.83E-111 | 1.61E-162 | WT |
| 2 | 16.9 | WT | 0.0027 | 0.0126 | 0.0156 | 1.06E-01 | 1.21E-31 | 1.75E-61 | WT |
| 3 | 15.9 | WT | 0.0086 | 0.0142 | 0.0182 | 5.07E-02 | 4.18E-04 | 2.90E-07 | WT |
| 4 | 15.9 | WT | 0.0052 | 0.0137 | 0.0152 | 3.58E-04 | 2.13E-47 | 1.60E-71 | WT |
| 5 | 13.4 | WT | 0.0043 | 0.0140 | 0.0168 | 2.83E-01 | 1.39E-18 | 9.77E-29 | WT |
| 6 | 14.6 | WT | 0.0039 | 0.0146 | 0.0164 | 1.65E-03 | 1.08E-65 | 1.72E-112 | WT |
| 7 | 16.4 | WT | 0.0030 | 0.0116 | 0.0148 | 1.47E-02 | 1.82E-55 | 1.58E-114 | WT |
| 8 | N/A | Delta | 0.0089 | 0.0033 | 0.0125 | 3.85E-27 | 3.26E-03 | 3.13E-99 | Delta |
| 9 | N/A | Delta | 0.0112 | 0.0018 | 0.0128 | 2.09E-30 | 4.97E-01 | 1.55E-69 | Delta |
| 10 | 19.4 | Omicron | 0.0135 | 0.0107 | 0.0046 | 2.82E-76 | 4.73E-82 | 9.54E-20 | Omicron |
| 11 | 19.7 | Omicron | 0.0113 | 0.0123 | 0.0042 | 5.38E-57 | 3.09E-77 | 1.22E-14 | Omicron |
| 12 | 19.7 | Omicron | 0.0113 | 0.0104 | 0.0057 | 2.60E-40 | 1.74E-40 | 9.31E-22 | Omicron |
| 13 | 19.3 | Omicron | 0.0139 | 0.0122 | 0.0019 | 1.11E-93 | 1.27E-123 | 4.25E-03 | Omicron |
| 14 | 18.6 | Omicron | 0.0124 | 0.0131 | 0.0038 | 6.03E-76 | 1.19E-106 | 4.05E-11 | Omicron |

**Extended Data Fig. 4: Kinetic barcoding measurement of clinical samples.**

a) Kernel function of all samples measured at assay endpoint.

b) Clinical sample information and the difference between the clinical sample versus the standard curves as determined by RMSE or the Kolmogorov–Smirnov test. The grey shading indicates incorrect predictions.

**Supplementary Video 1: Cas13a droplet assay workflow**

Automated droplet generation, incubation, cartridge loading, and wide-field microscope imaging of the Cas13a droplet assay.

**Supplementary Video 2: Molecular dynamics simulation of Cas13a in complex with 6T igRNA**

LbuCas13a (Cyan), 6T igRNA (Red) and target RNA (Purple) were simulated for 200 ns in an NVT ensemble. The HEPN domain is highlighted in yellow.

**Supplementary Video 3: Molecular dynamics simulation of Cas13a in complex with 12T igRNA**

LbuCas13a (Cyan), 12T igRNA (Red) and target RNA (Purple) were simulated for 200 ns in an NVT ensemble. The HEPN domain is highlighted in yellow.

**Supplementary Video 4: Molecular dynamics simulation of Cas13a in complex with 12A igRNA**

LbuCas13a (Cyan), 12A igRNA (Red) and target RNA (Purple) were simulated for 200 ns in an NVT ensemble. The HEPN domain is highlighted in yellow.

### Supplementary Table 1: Oligonucleotides used in this study

| Name | Sequence | Vendor | Used in |
|---|---|---|---|
| crRNA 2 | GACCACCCCAAAAAUGAAGGGGACUAAAAC**GGUCCACCAAACGUAAUGCG** | Synthego | Fig. 1G-H |
| crRNA 4 | GACCACCCCAAAAAUGAAGGGGACUAAAAC**UUUGCGGCCAAUGUUUGUAA** | Synthego | Fig. 1A-H, 2A-B, 3B, Ext Data Fig. 2A-B, 2E-I, 2K |
| crRNA 11 | UAGACCACCCCAAAAAUGAAGGGGACUAAAAC**GCUUGUGUUACAUUGUAUGC** | Synthego | Fig. 2A, 2C, Ext Data Fig. 2A-B, 2E-H |
| crRNA 12 | UAGACCACCCCAAAAAUGAAGGGGACUAAAAC**AAUUUGAUGGCACCUGUGUA** | Synthego | Fig. 2A, 2D, 2G-J, Ext Data Fig. 2A-H, 2J-K |
| HCoV crRNA 6 | GACCACCCCAAAAAUGAAGGGGACUAAAAC**CCUCUCUGGUAGGAACACGC** | Synthego | Fig. 2G-J |
| igRNA 4 (2T) | TTTTTTTTTT GACCACCCCAAAAAUGAAGGGGACUAAAAC**UUUGCGGCCAAUGUUUGUAA** | IDT | Fig. 3B-D, Ext Data Fig. 3 A-E |
| igRNA 4 (4T) | TTTTTTTTTTTC GACCACCCCAAAAAUGAAGGGGACUAAAAC**UUUGCGGCCAAUGUUUGUAA** | IDT | Fig. 3B |
| igRNA 4 (5T) | TTTTTTTCAGATC GACCACCCCAAAAAUGAAGGGGACUAAAAC**UUUGCGGCCAAUGUUUGUAA** | IDT | Ext Data Fig. 3B |
| igRNA 4 (6T) | TTTTTTTTTTTTTC GACCACCCCAAAAAUGAAGGGGACUAAAAC**UUUGCGGCCAAUGUUUGUAA** | IDT | Fig. 3B |
| igRNA 4 (7T) | TTTTTTTTTTTTTTT GACCACCCCAAAAAUGAAGGGGACUAAAAC**UUUGCGGCCAAUGUUUGUAA** | IDT | Ext Data Fig. 3A |
| igRNA 4 (8T) | TTTTTTTTTTTTTTTC GACCACCCCAAAAAUGAAGGGGACUAAAAC**UUUGCGGCCAAUGUUUGUAA** | IDT | Fig. 3B |
| igRNA 4 (10T) | TTTTTTTTTTTTTTTTTC GACCACCCCAAAAAUGAAGGGGACUAAAAC**UUUGCGGCCAAUGUUUGUAA** | IDT | Fig. 3B |
| igRNA 4 (12T) | TTTTTTTTTTTTTTTTTTTC GACCACCCCAAAAAUGAAGGGGACUAAAAC**UUUGCGGCCAAUGUUUGUAA** | IDT | Fig. 3B, Ext Data Fig. 3A |
| igRNA 4 (8A) | AAAAAAAAAACAGATC GACCACCCCAAAAAUGAAGGGGACUAAAAC**UUUGCGGCCAAUGUUUGUAA** | IDT | Fig. 4, Ext Data Fig. 3A, Ext Data Fig. 4 |
| igRNA 4 (12A) | AAAAAAAAAAAAAAAAAAAA GACCACCCCAAAAAUGAAGGGGACUAAAAC**UUUGCGGCCAAUGUUUGUAA** | IDT | Fig. 3B, Ext Data Fig. 3A |
| SC2 Delta igRNA (AT) | ATTTCAGATC GACCACCCCAAAAAUGAAGGGGACUAAAAC**AAUAAACUCCACUUUCCAUC** | IDT | Fig. 3C-D, Ext Data Fig. 3B-E |
| IAV igRNA (TTTT) | TTTTTTCAGATC GACCACCCCAAAAAUGAAGGGGACUAAAAC**CCCUAGUAGUUCAUAUAGGA** | IDT | Fig. 3C-D, Ext Data Fig. 3B-E |
| SC2 Delta igRNA (TT) | TTTTCAGATC GACCACCCCAAAAAUGAAGGGGACUAAAAC**AAUAAACUCCACUUUCCAUC** | IDT | Fig. 4, Ext Data Fig. 4 |
| SC2 omicron crRNA | GACCACCCCAAAAAUGAAGGGGACUAAAAC**UCGCGCCCCACCAUUCUGGU** | Synthego | Fig. 4, Ext Data Fig. 4 |
| crRNA4 20nt target | GCACAGCAGAAAATCTCTGC**UUACAAACAUUGGCCGCAAA**CCACAG | IDT | Ext Data Fig. 2K |
| crRNA12 20nt target | GCACAGCAGAAAATCTCTGC**UACACAGGUGCCAUCAAAUU**CCACAG | IDT | Ext Data Fig. 2K |
| PolyU Reporter | /ALEXA488/**UUUUU**/3IABKFQ/ | IDT | Fig. 1-4, Ext Data Fig. 1-4 |

DNA is underlined. The 20-nt oligonucleotides complementary to the crRNA spacer sequence is indicated in bold.
