## Supplemental Methods for "Programmable kinetic barcoding for multiplexed RNA detection with Cas13a"

Son, et al.

*Protein purification*

Protein purification was performed as previously described[1]. Briefly, the LbuCas13a expression vector contains the codon-optimized Cas13a genomic sequence, N-terminal His6-MBP-TEV cleavage site sequence, and a T7 promoter binding sequence (Addgene Plasmid #83482). The protein is expressed in Rosetta 2 (DE3) pLysS E. coli cells in Terrific broth at 16°C overnight. Soluble His6-MBP-TEV-Cas13a was isolated over metal ion affinity chromatography and the His6-MBP tag was cleaved with TEV protease at 4°C overnight. Cleaved Cas13a was loaded onto a HiTrap SP column (GE Healthcare) and eluted over a linear KCl (0.25-1.0M) gradient. Cas13a-containing fractions were further purified via size-exclusion chromatography on a S200 column (GE Healthcare) in gel filtration buffer (20 mM HEPES-K pH 7.0, 200 mM KCl, 10% glycerol, 1 mM TCEP) and were subsequently flash frozen for storage at -80°C.

*Preparation of SARS-CoV-2 RNA segments*

*In vitro* RNA transcription is performed as previously described[1]. SARS-CoV-2 N gene, S gene (WT), and S gene with D614G mutation were transcribed off a single-stranded DNA oligonucleotide template (IDT) using HiScribe T7 Quick High Yield RNA Synthesis Kit (NEB) following manufacturer's recommendations. Template DNA was removed by addition of DNase I (NEB), and IVT RNA was subsequently purified using RNA STAT-60 (AMSBIO) and the Direct-

Zol RNA MiniPrep Kit (Zymo Research). RNA concentration was quantified by Nanodrop and copy number was calculated using transcript length and concentration.

*Preparation of full viral genomic RNA*

Full viral genomic RNAs are purified as previously described[1]. Isolate USA-WA1/2020 of SARS-CoV-2 (NR-52281 BEI Resources) and CA427 (B.1.427, received from CA DPH) were propagated in Vero CCL-81 cells. Isolate Amsterdam I of HCoV-NL-63 (NR-470, BEI Resources) was propagated in Huh7.5.1-ACE2 cells (all virus cultures were performed in a Biosafety Level 3 laboratory). RNA was extracted from the viral supernatant via RNA STAT-60 (AMSBIO) and the Direct-Zol RNA MiniPrep Kit (Zymo Research).

*SARS-CoV-2 patient samples*

Patient samples were obtained from a UC Berkeley campus testing program, as previously described[2]. Briefly, human mid-turbinate nasal/oropharyngeal swabs were collected, processed, and tested by running qRT-PCR on extracted RNA using primers for SARS-CoV-2. Ct values were obtained using primers for the N gene, S gene, and Orf1ab of the SARS-CoV-2. Sequence data for each sample was used to identify SARS-CoV-2 variants. The samples used in this study were non-identifiable biospecimens, for which there was no link to identifiable subject information. Only secondary data analysis was carried out on the samples, and the research data will not be held for inspection by nor submitted to the FDA.

*crRNA design*

All unmodified crRNAs targeting SARS-CoV-2, SARS-CoV-2 variants Delta (SΔ157-158) and Omicron (NΔ31-33), HCoV-NL-63, or Influenza H3N2 were ordered as desalted oligonucleotides from Synthego at a scale of 5 nmole (Supplementary Table 1). Select crRNAs have also been previously described[1,3]. All DNA-modified crRNAs are ordered from Integrated DNA Technologies at a scale of 4 nmole (Supplementary Table 1).

*Bulk Cas13a nuclease assays*

LbuCas13a-crRNA RNP complexes were first preassembled at 133 nM equimolar concentration for 15 minutes at room temperature and then diluted to 25 nM LbuCas13a in cleavage buffer (20 mM HEPES-Na pH 6.8, 50 mM KCl, 5 mM MgCl2, and 5% glycerol) in the presence of 400 nM of reporter RNA (5'-Alexa488rUrUrUrUrU-IowaBlack FQ-3'), 1 U/µL Murine RNase Inhibitor (NEB, Cat# M0314), 0.1 vol% IGEPAL 630 (Fisher, Cat# ICN19859650), and varying amounts of target RNA. For reactions using more than one crRNA, multiple crRNAs were combined at an equal concentration and subsequently the total crRNA mix was assembled with Cas13a at 133nM equimolar concentration. 25nM of RNP complex is used unless specified otherwise. The reaction mix is measured either in bulk or in droplet following emulsification (see *Droplet formation*). For the bulk Cas13a assay, the reactions were loaded into a 384-well (Corning, Cat #3820) incubated in a fluorescence plate reader (TECAN, infinite 200Pro) for 60 minutes at 37°C with fluorescence measurements taken every 2.5 minutes ($\lambda_{ex}$:485 nM; $\lambda_{em}$:535 nM; Gain: 130). Background-corrected fluorescence values were obtained by subtracting fluorescence values obtained from reactions carried out containing only reporter and buffer. The slope of Cas13a reaction is calculated by simple linear regression of raw data from time 0 for the set duration.

*Droplet formation*

To emulsify the Cas13a reaction mix, 20 µL of the aqueous mix was combined with 100 µL of HFE-7500 oil supplemented with 2% (w/w) PEG-PFPE amphiphilic block copolymer surfactant (008-Fluoro-surfactant, RAN Biotechnologies) in a 0.2 mL 8 tube-strip. The oil/aqueous mix was emulsified by repeated, fully-automated pipetting using an electronic 8-channel pipette (Integra-biosciences, Part #4623) fitted with a 300 µL pipet tip (Integra-biosciences, Part #3835). A sample volume of 110 µL was mixed for 150 repetitions at the maximum speed (speed 10) to generate droplets of a narrow size range. The emulsion was either directly loaded into a flow cell for the time course imaging or incubated in a heating block at 37˚C before being transferred and imaged. In cases where imaging could not be performed immediately after the reaction, the emulsion was quenched on ice until imaging. For the time course imaging experiment, the emulsion was quickly separated by spinning in a speed-controlled mini-centrifuge (~50 rpm) for 10 seconds, the oil was completely removed from the bottom of the tube, and the emulsion was transferred into a custom imaging chamber after several cycles of gentle manual mixing. It took approximately 10 minutes from droplet formation to the start of image acquisition.

*A custom imaging chamber for droplet imaging*

The sample flow cell was prepared by sandwiching double-sided tape (~20µm thick, 3M Cat# 9457) between an acrylic slide (75mm x 25mm x 2mm, laser cut from a 2mm-thick acrylic plate) and a siliconized coverslip (22mm x 22mm x 0.22mm, Hampton research Cat# 500829). Both surfaces are hydrophobic, promoting thin layers of oil between droplets and the surface. Siliconized coverslips were rinsed with isopropanol to remove any auto-fluorescent debris (20 minutes sonication) and spin dried prior to assembly. 15 µL of sample emulsion is loaded into the flow cell by capillary action, after which the inlet and outlet are sealed with Valap (1:1:1 vasoline:lanoline:paraffin).

For an increased throughput of samples using the pipetting robot, a custom imaging chamber was built containing 8 independent flow cells. Each flow cell (39mm x 6.6mm x 0.026mm) consisted of a double-sided adhesive (ARcare 8026, Adhesives Research) bonded between an acrylic slide (75mm x 50mm x 1.5mm, laser cut from a 1.5mm-thick acrylic plate) and a large glass microscope slide (75mm x 51mm x 1.2mm). Prior to the assembly, the large glass microscope slides were sonicated in 2% liquid detergent (Neutrad, Decon labs) for 30 minutes at 55˚C followed by a rinsing step with MilliQ water and blow dry using a compressed air blow gun. After the assembly, the imaging chamber was placed on an acrylic chamber holder where the pipetting robot loaded the sample emulsion on the inlet ports of each well simultaneously using an automated program. Once the droplets filled the total volume of the independent flow cells, the imaging chamber was ready to be imaged.

*Microscopy and data acquisition*

Droplet imaging was carried out for both high resolution (low throughput) and low resolution (high throughput) studies. The high-resolution imaging was carried out on an inverted Nikon Eclipse Ti microscope (Nikon Instruments) equipped with a Yokogawa CSU-X spinning disk. A 488-nm solid state laser (ILE-400 multimode fiber with BCU, Andor Technologies) was used to excite the RNA probe, and the fluorescence light was spectrally filtered with a 535/40nm emission filter (Chroma Technology) and imaged on an sCMOS camera (Zyla 4.2, Andor Technologies). A 20x water-immersion objective (CFI Apo LWD Lambda S, NA 0.95) was used with the Perfect Focus System to monitor droplets during the reaction or to accurately quantify fluorescence signal at reaction endpoint. 400nM of fluorescent reporter was added to the reaction. Images were acquired through Micro-Manager with 500ms exposure time and 2x2 camera binning. Typically, 16 field-of-views (FOV) were acquired every 30 seconds for the time course imaging and 36 FOVs are acquired for the endpoint imaging. The low-resolution imaging was carried out on a Zeiss Axiovert with a

2.5x objective (Plan-NEOFLUAR, NA 0.085) to capture droplet reaction endpoints after 30 minutes or 1hr of reaction time. A Xenon light source with a 470/40 excitation filter (Chroma Technology) and 535/40nm emission filter (Chroma Technology) were used. 1.6μM of fluorescent reporter was added to the reaction, which increased both the signal and background compared to the reaction containing 400nM reporter. To image all the droplets for 8 independent reactions simultaneously on the custom imaging chamber, 7 FOVs were acquired per reaction with 6s exposure time without camera binning.

*Image analysis – droplet detection*

We used MATLAB (Mathworks R2020b) to detect positive droplets and quantify fluorescence signals from microscopy images. First, the grayscale images were converted to binary images based on a locally adaptive threshold. The threshold is defined generously to select all the positive droplets and potentially some negative droplets or debris at this stage. Second, connected droplets were separated by watershed transform. Third, individual droplets are identified by looking for circular continuous regions and droplet parameters such as radius, circularity, and fluorescence signal are quantified. Fluorescence signal is quantified in two different ways: the mean fluorescence signal of a droplet reflecting the density of cleaved reporter; the total fluorescence signal reflecting the total amount of cleaved reporter within a droplet. Lastly, positive droplets are chosen based on their circularity and total fluorescence signal by applying a threshold that was consistently used throughout the experiments.

*Image analysis – droplet tracking in time course images*

To quantify signal accumulation in the same droplet over time, we associated droplets over time based on their motion estimated by a Kalman filter in MATLAB. The filter is used to predict the track's location in each frame and determine the likelihood of each detection being assigned to each track. Only the droplets showing continuous trajectories in time and magnitude are selected for the downstream analysis.

*Comparison of single Cas13a reaction with enzyme kinetics*

We analyzed the Cas13a reaction with a single crRNA (Fig. 1E) using the Michaelis-Menten enzyme kinetics model with the quasi-steady-state approximation:

$$v = \frac{K_{cat}[E_0][S]}{K_M + [S]}$$

where $v$ is the reaction rate, $[E_0]$ is ternary Cas13a, $[S]$ is the free RNA reporter, $K_{cat}$ and $K_M$ are the catalytic rate constant and the Michaelis constant. We assumed low substrate concentration ($[S]$ << $K_M$) since the RNA reporter $[S]$ is 400nM and $K_M$ is estimated to be larger than 1μM[4] and simplified the equation to:

$$v = \frac{K_{cat}[E_0][S]}{K_M}$$

$v/[E_0]$ is turnover frequency, or the reciprocal of the mean waiting time <1/t> in the single-molecule Michaelis-Menten framework[5], and it can be obtained from Fig. 1D and E after converting the fluorescence signal to molar concentration of cleaved reporter based on a calibration.

*Data analysis – Cas13a time trajectories*

We processed the raw signal in a series of steps prior to analysis: First, we corrected for the global signal fluctuation, which arise from a slight drift in z-focus even with the Perfect Focus System; we characterized the global signal from the background droplets as the mean of their pixel values and divided it from the positive droplet signal in each image frame. Second, we corrected for the photobleaching; we first characterized the signal decay rate from >200 Cas13a curves exhibiting negative slopes and positive initial signals. We modeled photobleaching as a linear function of the initial signal based on the observed linear relationship between the decay rate versus initial signal ($R^2$ = 0.87). Using the model, we corrected for photobleaching in each trajectory point-by-point. Third, we filtered the trajectories with a weak Savitzky-Golay filter (order 5, frame length 9) to remove the high frequency measurement noise while preserving overall structure of the curve. Lastly, we calculated instantaneous slopes by dividing signal change between frames by the frame interval and removed single outliers exhibiting high positive or negative slopes. To characterize key parameters of Cas13a kinetics, we analyzed individual trajectories in two different domains: First, we determined the slope, $T_{init}$, and RMSD from signal time trajectories by linear regression. Since $T_{init}$ indicates time since the droplet reaction was started, we added a constant time (12.5 minutes) that took from Cas13 droplet formation until the beginning of time course imaging. Second, we determined the $slope_{fast}$, $slope_{slow}$, and a fraction spent in each period by fitting a Gaussian pdf to the instantaneous slope distribution. We compared the model qualities between the single versus binary Gaussian pdfs using Akaike's Information Criterion (AIC) to determine whether a trajectory exhibits two different periods of slope of not.

*Data analysis – kinetic barcoding with unmodified crRNAs (corresponding to Fig. 2)*

We used the slope and RMSD of individual signal trajectories to compare Cas13a reactions between different target-crRNAs. We first performed binary classification of trajectories based on supported vector machine (SVM) in MATLAB. For this, we collected 200 to 400 signal trajectories in each condition, in two or more independent experiments per condition to prevent bias. We converted the trajectories into a 2D array consisting of the slope and RMSD and divided the array into a training and a validation set. We then trained an algorithm using the training set with the known answers (i.e. target-crRNA condition) and classified the validation set. The accuracy of identifying individual trajectories was 75% for HCoV-NL-63 vs SARS-CoV-2. To assess the difference between two groups of trajectories, we evaluated p-values for the predicted classes using a two-tailed Student's t-test.

*Data analysis – kinetic barcoding with signal slope of IgRNAs (corresponding to Fig. 3)*

The goal of this analysis was to quantify virus compositions of a sample from its slope distribution, assuming any permutations of single or multiple co-infections could be present. The slopes obtained from either unmodified crRNA or IgRNAs exhibited distributions with partial overlap between each other. In order to statistically predict the class of a measured slope, we used a multiclass error-correcting output codes (ECOC) model using support vector machine (SVM) binary learners. We trained a model based on the slope distribution of 4 different classes (i.e. crRNA and IgRNAs). From the model, we obtained class posterior probabilities, which assigns relative probabilities that a given slope could be measured from one class versus the others. To quantify the virus composition of an unknown sample, we converted the slope distribution to the matrix of class posterior probabilities. We then quantified virus compositions by summing the posterior probability of individual classes.

*Data analysis – kinetic barcoding with endpoint signal of IgRNAs (corresponding to Fig. 4)*

The goal of this analysis was to classify the variant information of a sample from three possible cases (i.e. SC2 wildtype, SC2 delta, SC2 omicron). For this, we characterized the kernel function of individual SARS-CoV-2 variants by measuring fluorescence signal with virus RNA isolated from a cell. For a patient sample, we compared its kernel function with the three reference functions and quantified their difference in terms of RMSE. We chose the variant with the lowest RMSE value. As an alternative to using the kernel function, we also used Kolmogorov–Smirnov test, which compares two cumulative distribution functions (CDF) without any filters and obtained consistent result.

*Molecular dynamics simulations*

Simulations were based on the structure of *Leptotrichia buccalis* (Lbu) Cas13a in complex with a crRNA and target RNA (PDB: 5XWP), obtained by single-wavelength anomalous diffraction at 3.08 Å resolution[6]. The complete structure with inclusion of missing atoms and reinstated catalytic residues R1048 and H1053 which were mutated for alanine in the crystal structure, were obtained from the SWISS-MODEL homology modelling repository[7,8]. Pymol was used to mutate RNA sequences and build nucleotides for the inhibitory segment. All histidine residues were treated as singly protonated in the ε position based on previous literature on Cas13a simulations[9].

All MD simulations were conducted using GROMACS (version 2024.1)[10] and the AMBER ff14SB force field[11], including the χOL3 corrections for RNA[12,13] and OL15 corrections for DNA [14–16]. We employed the TIP3P model for water molecules[17].

Systems were solvated on a dodecahedron box with 50 mM KCl, 5 mM $MgCl_2$ and neutralized by addition of $Na^+$ counterions to simulate reaction conditions. Firstly, all systems were subjected to a 50000-step energy minimization using the steepest descent algorithm to prevent any steric clashes. Then, the systems were heated in two consecutive isochoric (NVT) ensembles, to 300K for 100 ps and then to 310K for 100 ps. Temperature was kept constant using velocity rescaling with a 0.1 ps time constant. Afterwards, the system’s density was equilibrated in the isobaric ensemble (NPT) for 100 ps by coupling the pressure to 1 atm using a C-rescale barostat with time constant of 2.0 ps. All bond lengths of hydrogen atoms were constrained using the SHAKE algorithm. Long range electrostatic calculations were conducted using the particle-mesh-Ewald algorithm. Electrostatic and Van der Waals cutoff values were both set to 1.2 nm. Finally, all position constraints were released, and production runs of 200 ns were carried out in the NVT ensemble using a 2-fs integration step.

Distances for Fig. 3D and 3E were computed by measuring the shortest distance from any atom of the DNA effector region to the center of mass for the catalytic site (R472, H477, R1048, H1053) across the entire simulation time.

METHOD REFERENCES
